# Development of a duplex real-time PCR assay for the differentiation of *Blastomyces dermatitidis* and *B. gilchristii* and a retrospective study of blastomycosis in New York (2005-2019)

**DOI:** 10.1101/2020.08.11.20172932

**Authors:** Mitchell Kaplan, YanChun Zhu, Julianne V Kus, Lisa McTaggart, Vishnu Chaturvedi, Sudha Chaturvedi

**Author notes:** Corresponding author: Sudha Chaturvedi. Mitchell Kaplan and YanChun Zhu contributed equally.

## Abstract

Blastomycosis due to *Blastomyces dermatitidis* and *B. gilchristii* is a notable cause of respiratory mycoses in North America with recurrent outbreaks. We developed a highly sensitive, specific, and reproducible Taqman duplex real-time PCR assay for the differentiation of *B. dermatitidis* and *B. gilchristii*. The new assay permitted retrospective analysis of *Blastomyces* cultures (2005 to 2019), and primary clinical specimens (2013-2019) from NY patients. *Blastomyces dermatitidis* was the causal agent for the majority of 38 cases while *B. gilchristii* was implicated in five cases; a rare finding reported from New York. The duplex real-time PCR assay will be useful for further understanding of ecology and epidemiology of blastomycosis caused by *B. dermatitidis* and *B. gilchristii*.

## INTRODUCTION

For the longest time, *Blastomyces dermatitidis* was considered a single species within the *Blastomyces* genus as the etiologic agent of pulmonary and disseminated blastomycosis (1). This knowledge was used to develop several diagnostic tests. With the advancement of molecular tools, several distinct genetic populations were recognized within the *Blastomyces* genus, and that led to the creation of new species including *B. gilchristii* (a cryptic species phenotypically indistinguishable from *B. dermatitidis)* (2, 3), *B. percursus*, and *B. silverae* (4, 5). A rare cause of blastomycosis from *B. helicus* in western part of North America and *B. percursus* in Africa was also reported (6, 7). Additionally, *Emmonsia parva* (now *Blastomyces parvus)* and *Emmonsia helica* (now *Blastomyces helicus)* were assigned to the *Blastomyces* genus (5). *Blastomyces dermatitidis* is the predominant species in North America, but *B. gilchristii* predominates in the hyperendemic regions of Northwestern Ontario and northern Wisconsin including large outbreaks (6). The ecological niche (s) of these two closely related species remains undefined. Similarly, the role of remaining rare *Blastomyces* species in the overall incidence and outcome of blastomycosis cases requires further investigations.

All *Blastomyces* spp. are thermally dimorphic fungi, exist as a mold form at ambient temperature in the environment but convert to the pathogenic yeast form *in vitro* at 37°C or when susceptible mammalian host inhales the conidia of the mold form. The yeast form of these pathogens can be microscopically differentiated with *B. dermatitidis* and *B. gilchrtstii* producing abundant large (8-20 mm) broad-based budding yeasts; *B. percursus* producing large yeast-like cells from fragmented swollen hyphal cells and *B. helicus* producing variably shaped yeast cells (4 to 5 μm) in short chain, while thin walled giant cells and occasional broad-based budding yeast like-cells are seen in *B. parva* and *B. silverae* (8). The distinct yeast morphology might be helpful in the species differentiation from the primary specimens; but this approach would require significant mycology expertise, which is not always available in diagnostic laboratories. Although culture and histopathology are useful techniques in the diagnosis of blastomycosis, the importance of molecular methods cannot be emphasized enough for the accurate identification of newer *Blastomyces* species. There are currently no molecular or serologic diagnostic tests for rapid and accurate identification of different species within *Blastomyces* genus. An available commercial test (Gen-Probe, Inc., San Diego, CA) is rather limited as it can be used only with pure cultures of *B. dermatitidis*. Previously, we developed a rapid, and specific real-time PCR assay for the identification and detection of four known haplotypes of *B. dermatitidis* because at the time methods for distinct species level determination of *Blastomyces* genus were not available (9).

Outdoor exposure and proximity to water have been associated with blastomycosis (10). Blastomycosis is reportable in only five states in the US, namely Arkansas, Louisiana, Michigan, Minnesota, and Wisconsin (6). In New York State, Blastomycosis is considered an emerging disease [http://www.nycasm.org/Alerts/Notification_101674.pdf]. Despite several blastomycosis cases reported from NY in the last decade, the disease endemicity is difficult to discern as it is not reportable in NY (11-14). Likewise, in the absence of a skin test, it is difficult to assess the geographic distribution of the exposure. Limited canine blastomycosis data suggest that it is endemic in the Saint Lawrence River Valleys on the New York-Canada border (11). While pneumonia is the most common manifestation of blastomycosis, approximately half of all cases are asymptomatic (15). Infection however can lead to severe and fatal disease, often as a result of respiratory failure (16). Disseminated infection can involve any organ, often including cutaneous abscesses and osteomyelitis, and is frequently accompanied by fever, weight loss, and night sweats (17).

The BAD-1 is an important conserved adhesion-promoting protein and virulence factor of *B. dermatitidis* (18). Two large insertions were noted in the *B. gilchristii BAD1* (19), while this gene was absent in African strains of *Blastomyces* spp. (20). Several conventional PCR assays were designed using *BAD1* gene (21) for the detection of *B. dermatitidis* DNA from clinical and soil samples (22, 23). Sidamonidze et al. (2012) developed a real-time PCR assay targeting *BAD1* gene to identify *B. dermatitidis* in culture and primary specimens. In the present investigation, we exploited the sequence variations observed within *BAD1* gene to develop a duplex real-time PCR assay for the differentiation of *B. dermatitidis* and *B. gilchristii*. We present data on the sensitivity, specificity, and reproducibility of the assay and retrospective analysis of blastomycosis cases from 2005 to 2019 in New York. Our results highlight the preponderance of *B. dermatitidis* and a minor population of *B. gilchristii* in New York cases.

## MATERIALS AND METODS

### Strains, primary specimens, and DNA

Seventy-nine isolates of *Blastomyces* spp. were evaluated in this study. These included 48 clinical isolates from the sporadic cases of blastomycosis obtained between 2005 to 2019 from New York; canine (6), feline (1), sea lion (2), bat (1), polar bear (1), soil (4), human (2) obtained from Dr. Gene M. Scalrone, Dept. of Biological Sciences, Idaho State University; American Type Culture Collection (ATCC) strains MYA2586 (soil), MYA2585 (dog), 56214 (human, Africa), and 56216 (human, Africa) and five each well characterized strains of *B. dermatitidis* and *B. gilchristii* from Public Health Ontario Laboratory, Toronto, Canada were part of this investigation. A total of 33 primary clinical specimens from New York patients including 13 paraffin tissues, 6 bronchial washes, 5 lung tissues, 3 cerebrospinal fluids, 2 skin lesion tissues, 2 bone marrows, one each blood, brain lesion tissue, and sputum submitted for *B. dermatitidis* identification, was also part of this investigation. Additionally, DNA from closely and distantly related fungal pathogens were included in this investigation (9).

### DNA extraction

DNA from *Blastomyces* spp. was extracted using Qiagen DNA mini kit on the Qiacube automated extractor. In brief, fungal growth (approximately 5×5 mm in size) was removed from the agar surface using a sterile loop and added into a lysing reagent in the BSL3 laboratory. The fungal suspension was incubated at 90°C for one hour and then brought to the BSL2 laboratory for further processing. The heat-killed fungal suspension was homogenized in the Precellys homogenizer at 6,500 RPM for 15 second each at three times (Program #5; 6500-3×60-015). The homogenized suspension was transferred to a 2 ml screw cap tube leaving behind the beads. DNA from samples were extracted using the Qiagen DNA mini kit in the QiaCube semi-automated DNA extractor resulting in 50 μL of eluted DNA. All extracted DNAs were stored at −80°C. DNA of fungal species (yeasts and molds) other than *Blastomyces* spp. were procured from Wadsworth Center Mycology Laboragtory (WCML) DNA Collection Repository.

DNA from paraffin fixed tissue was first sectioned by the Wadsworth Center Histopathology Core, then using 1 mL of xylene, the paraffin was dissolved, followed by two washes using 1 mL of 100% ethanol. The sample was then dried and extracted using the Qiagen DNA mini kit as described above with an incubation temperature of 70°C instead of 90°C. For non-fixed tissues, approximately 5×5 mm of tissue was used and extraction was also done using the Qiagen DNA mini kit as described for the isolates with a incubation temperature of 70°C instead of 90°C.

DNA from soil samples were manually extracted using the DNeasy PowerSoil Kit as described previously (24). In brief, about 0.25 g of soil sample was added to the Powerbead tube with 60 μL C1 solution followed by homogenizing in a vortex genie for 20 minutes. Sample was then processed through multiple spin column washes and DNA was eluted in 100 μL of elution buffer.

#### Primers and probes

Primers and probes for the real-time PCR assay were designed from the promoter region of the *BAD1* gene using Geneious R9 9.1.6 software. The choice of *BAD1* promoter was based upon our earlier successful utilization for this region for *B. dermatitidis* identification by real-time PCR assay (9). Multiple alignment of*BAD1* promoter sequences revealed unique sequences for *B. dermatitidis* and *B. gilchristii*, which were used for primers and probes design (Supplementary Figure 1). Oligonucleotide sequences of the primers and probes for *B. gilchristii*, and *B. dermatitidis* are as follows: V2556 *(Bg;* forward) 5’-ATGGGTGCAAAATCCGCCTA-3’, V2558 (Bg; reverse) 5’AATCTAGAAGCTGGAGCGCC -3’, V2557 *(Bg;* probe) 5’-/6-FAM- CCGTACTCCCTCCCCGGTACTCC-ZEN/3’ IB FQ. V2559 (Bd; forward) 5’- GCAAAATCCGCCTACTACTA-3’, V2561 *(Bd;* reverse) 5’ AGCTGAACCTGGAAGTATTG -3’, V2560 *(Bd;* probe) 5’-/6-Cy5- TCCCTACCCCTGGCTACTTTTCT-TAO/3’ IB FQ.

#### Real-time PCR Assay

Each sample DNA (isolate or primary specimen) was tested in duplicate in 20 μl reaction volumes using an optical 96-well plate. Each reaction mixture contained 1× PerfeCTa Multiplex qPCR ToughMix (Quanta Biosciences), a 1,000 nM concentration of each *B. gilchristii* (V2556 & V2558) and *B. dermatitidis* (V2559 & V2561) primers, a 250 nM concentration of each *B. gilchristii* (V2557) and *B. dermatitidis* (V2560) probe, and 2 μl of DNA extracted from *Blastomyces* spp. or 5 μl of DNA extracted from primary clinical specimens. Each PCR run also included 2 μl of positive extraction control (M808; *B. dermatitidis)*, 2 μl of positive amplification controls (M808; *B. dermatitidis* and MB97; *B. gilchristii)* and 2 μl of negative extraction (extraction reagents only) and negative amplification (sterilized nuclease-free water) controls. To prevent any cross-contamination, a unidirectional workflow was followed by keeping reagent preparation, specimen preparation, and amplification/detection areas separate. Cycling conditions on the ABI 7500 FAST were initial denaturation at 95°C for 20 s, followed by 45 cycles of 95°C for 3 s and 60°C for 30 s. Based on limit of detection (LOD), a cycle threshold (*Ct*) value of ≤38 was reported as positive and >38 was reported as negative. Specimens were reported as inconclusive if PCR inhibition was observed for the primary specimens.

## RESULTS

### Assay sensitivity, specificity, and reproducibility-

The duplex real-time PCR assay was highly sensitive with linearity over five logs, a correlation coefficient of >0.99, and amplification efficiency of >92%. The limit of detection (LOD) of the assay was one pg gDNA per PCR reaction, which was within the linear range of the standard curve for *B. dermatitidis* and *B. gilchristii* (Fig. 1 and Table 1). These results indicated that the duplex real-time PCR assay targeting the *BAD1* gene is highly sensitive for the culture identification of *B. dermatitidis* and *B. gilchristii*. The assay was highly specific as it did not cross-react with other closely and distantly related fungal pathogens (Supplementary Table 2). Ten blinded genomic DNA received from Ontario, Canada, including five each of *B. dermatitidis* and *B. gilchristii*, identified both species accurately, further confirming the assay validity (Table 2). Additionally, DNA from *B. dermatitidis* and *B. gilchristii* strains run on three different days and within the same day in triplicates yielded consistent Ct values with cofficient of variance <5.0, confirming assay reproducibility (Supplementary Table 3 A and 3 B).

**Figure 1.**
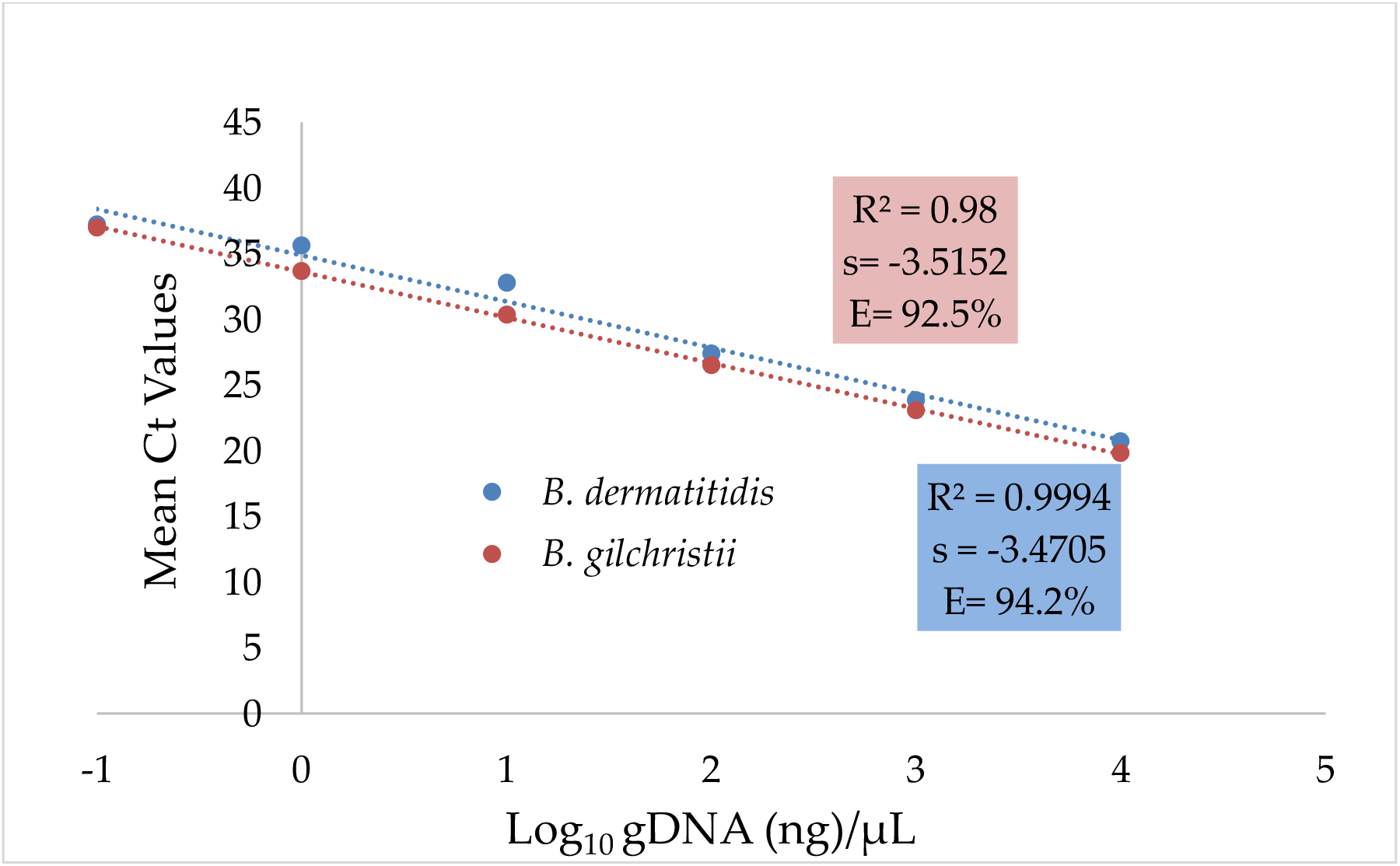
Sensitivity of duplex real-time PCR assay. Serial 10-fold dilution series of gDNA *of B. dermatitidis* and *B. gilchristii* were prepared and PCR was run in duplicate. The slope (s) and correlation coefficient (*R*) are shown.

**Table 1.**
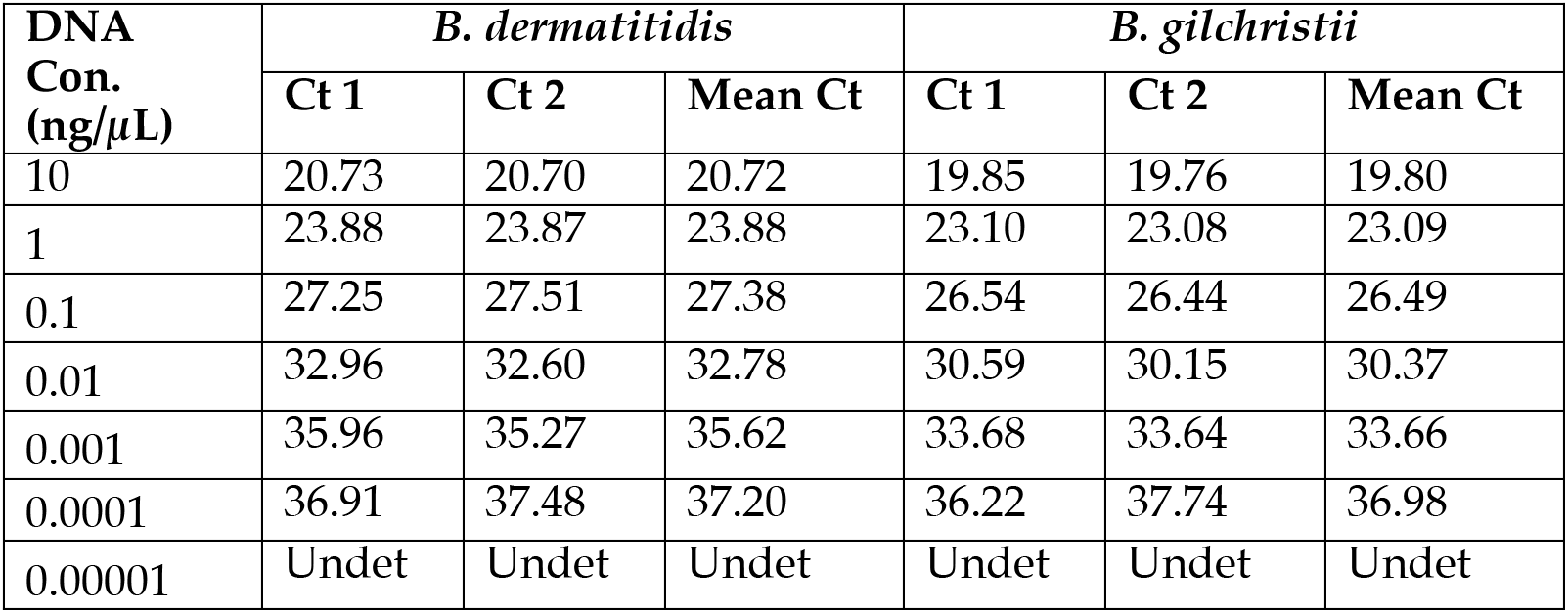
Sensitivity study of *B. dermatitidis* and *B. gilchristii*

**Table 2.** Blinded panel of DNA of *B. dermatitidis* and *B. gilchristii\**

| Sample No. | Single plex real-time PCR Assay Mean Ct | Duplex real-timePCR Assay |  | Final ID |
| --- | --- | --- | --- | --- |
|  |  | *B. dermatitidis* Mean Ct (Cy5) | *B. gilchristii* Mean Ct (FAM) |  |
| 1 | 23.02 | 23.48 | Undet | *B. dermatitidis* |
| 2 | 22.4 | Undet | 21.74 | *B. gilchristii* |
| 3 | 18.52 | 20.04 | Undet | *B. dermatitidis* |
| 4 | 22.31 | 22.64 | Undet | *B. dermatitidis* |
| 5 | 19.86 | Undet | 19.35 | *B. gilchristii* |
| 6 | 22.12 | Undet | 21.54 | *B. gilchristii* |
| 7 | 23.28 | Undet | 22.46 | *B. gilchristii* |
| 8 | 19.75 | 20.15 | Undet | *B. dermatitidis* |
| 9 | 22.23 | 22.39 | Undet | *B. dermatitidis* |
| 10 | 21.45 | Undet | 20.39 | *B. gilchristii* |

\*DNA was provided by Dr. Julianne V. Kus, University of Toronto, Canada, in a blinded fashion

### Retrospective analysis of *Blastomyces* isolates and primary specimens

The retrospective analysis of 79 isolates of *Blastomyces* spp. revealed 62 to be *B. dermatitidis*, and 15 to be *B. gilchristii* (Table 3 A). One isolate (ATCC 56214), which was identified by an older single-plex real-time PCR assay as *Blastomyces* spp., was neither *B. dermatitidis* nor *B. gilchristii* by the new duplex real-time PCR assay. It was later confirmed to be *B. percursus* by sequencing of the ribosomal genes and BLAST search. One strain of *B. emazntsi* (ATCC 21516), identified by sequencing, was neither picked up by the old single-plex real-time PCR nor by the new duplex real-time PCR assay further confirming the specificity of the duplex real-time PCR assay. Of 15 *B. gilchristii* isolates identified, 5 were well-characterized strains of *B. gilchristii* from Canada, 4 were from the Eagle River Wisconsin outbreak involving human, dog, and soil, and 6 were from five patients from NY. All six NY isolates of *B. gilchristii* confirmed by duplex real-time PCR assay, were also confirmed by sequencing of the *BAD1* and ribosomal genes (GenBank Accession No for *BAD1:* MT822768 - MT822773; GenBank Accession Number for *ITS:* MT822762 - MT822767 for *ITS)*. Of 62 *B. dermatitidis* isolates identified, 5 were well-characterized strains of *B. dermatitidis* from Canada, 15 were from Illinois, Kentucky, Minnesota, Tennessee, and Wisconsin involving animals or soil, and 42 isolates were from 38 patients from NY collected from 2005 to 2019. Of 33 primary specimens analyzed from 2013 to 2019, five specimens (one skin lesion, one bronchial wash, two tissue blocks of nasal mass, and one tissue block of unknown origin) from five patients were positive for *B. dermatitidis* DNA while none were positive for *B. gilchristii* DNA (Table 3 B).

**Table 3 A.**
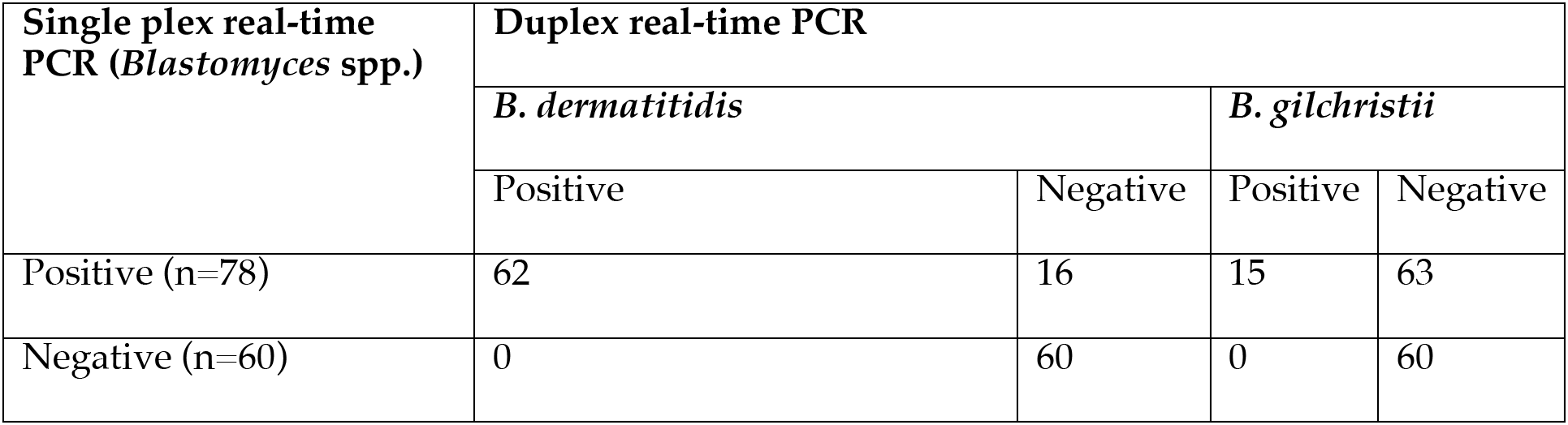
Validity of duplex real-time PCR assay for culture identification of *Blastomyces* spp.

**Table 3 B.** Validity of duplex real-time PCR assay for the direct detection of *Blastomyces* spp. DNA from primary specimens

| Single plex real-time PCR (*Blastomyces* spp.) | Duplex real-time PCR |  |  |  |
| --- | --- | --- | --- | --- |
|  | *B. dermatitidis* |  | *B. gilchristii* |  |
|  | Positive | Negative | Positive | Negative |
| Positive (n=5) | 5 | 0 | 0 | 5 |
| Negative (n=28) | 0 | 28 | 0 | 28 |

### Blastomycosis in New York

Analysis of 43 cases of blastomycosis from NY (Supplementary Table 1) revealed one to four cases per year from 2005 until 2016, but that number increased to 7 in 2017, and cases remained high with 9 identified in 2018 and 8 identified in 2019 (Fig. 2). Men were more frequently infected than women with *B. dermatitidis*, while number of cases was too small to derive this conclusion for *B. gilchristii*. Older people (over age 40) were found to be more prone to symptomatic infection irrespective of *B. dermatitidis* or *B. gilchristii. B. dermatitidis* was most commonly isolated from respiratory specimens, followed by skin/wounds/subcutaneous tissue, and bone, while all six isolates of *B. gilchristii* were isolated from respiratory specimens (Table 4). Geographic distribution of blastomycosis revealed that the majority of cases presented with symptoms lived in Mohawk Valley, Capital District and Finger Lakes. Few cases were also reported from other regions in NY (Supplementary Figure 2).

**Figure. 2.**
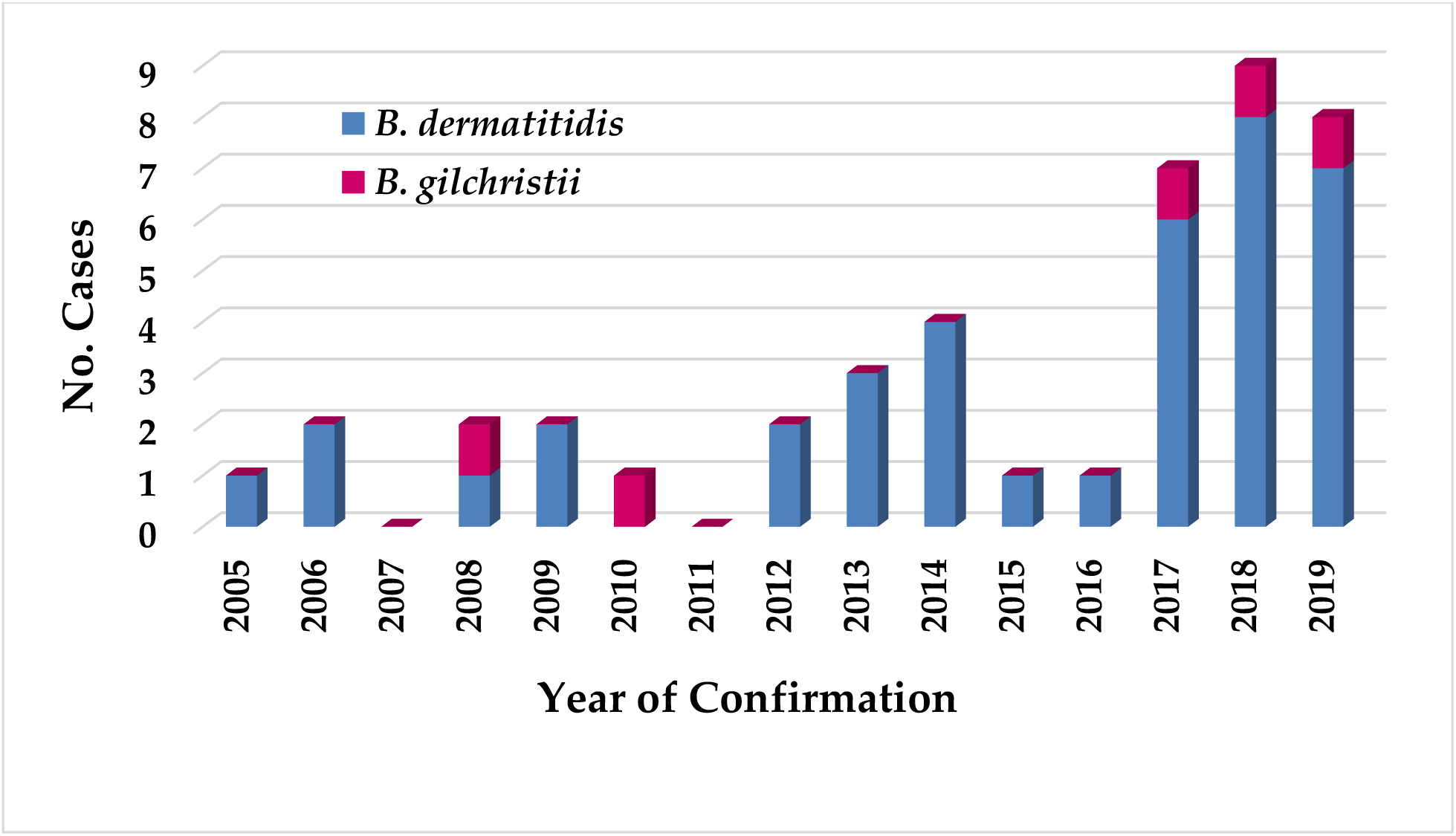
Blastomycosis cases in New York from 2005 to 2019. Blastomycosis cases earlier confirmed by single plex real-time PCR assay for isolates and primary specimens were analyzed retrospectively by the newly developed duplex real-time PCR assay. Of 43 cases, 38 were confirmed as *B. dermatitidis* and 5 as *B. gilchristii*. There was marked increase in blastomycosis cases from 2017 onwards.

**Table 4.** Characteristics of Mycology Laboratory confirmed blastomycosis cases in New York from 2005 to 2019

| Characteristic | Blastomycosis | *B. dermatitidis* | *B. gilchristii* |
| --- | --- | --- | --- |
| Patient Sex | n = 43 | n = 38 | n = 5 |
| M | 29 | 26 | 3 |
| F | 14 | 12 | 2 |
| Patient Age (Y) | n = 43 |  |  |
| 20 - 29 | 4 | 4 | 0 |
| 30-39 | 4 | 4 | 0 |
| 40-49 | 8 | 7 | 1 |
| 50-59 | 11 | 10 | 1 |
| 60-69 | 13 | 10 | 3 |
| >69 | 3 | 3 | 0 |
| Source of Specimen Isolation | n = 48 |  |  |
| Bronchial wash | 28 | 22 | 6 |
| Skin, wound, subcutaneous tissue | 15 | 15 |  |
| Bone | 4 | 4 |  |
| Brain | 1 | 1 |  |
| Specimen positive for DNA | 5 |  |  |
| Bronchial wash | 1 | 1 |  |
| Tissue block - Nasal mass | 2 | 2 |  |
| Tissue block-source not provided | 1 | 1 |  |
| Skin wound fluid | 1 | 1 |  |

### Analysis of soil samples

Two field studies were conducted to find the environmental source of *B. dermatitidis* and *B. gilchristii*. The first extensive collection focused on a variety of sites in and around the city of Amsterdam (Mohawk Valley), NY where a number of blastomycosis case patients lived or visited. The second more limited sampling took place in the Lake George (Capital District), NY area where one patient of blastomycosis resided. None of the sample yielded any positive DNA for *Blastomyces* spp. by our earlier developed single-plex real-time PCR assay (9) or newly developed duplex real-time PCR assay. None of the soil samples yielded any positive growth of *Blastomyces* spp. in culture (details not shown).

## DISCUSSION

In this study, we have developed a duplex real-time PCR assay to differentiate *B. dermatitidis* from *B. gilchristii* as these two closely related species possibly overlap in their endemicity in North America. The duplex real-time PCR assay targeting the putative promoter region of the *BAD1* gene was highly sensitive, specific, and reproducible. *BAD1* gene and its promoter have been extensively used for the differentiation of *Blastomyces* species using restriction fragment length polymorphism (RFLP), PCR and real-time PCR assays (9, 19, 23). The polymorphism in *BAD1* gene and markedly different sizes including 363-bp in *B. dermatitidis*, 663-bp in *B. gilchrstii* and absence of this gene in African isolates of *Blastomyces* (8, 9, 19, 20), proved this target as highly specific for the identification of *B. dermatitidis*, and *B. gilchristii* in the present investigation.

The retrospective analysis of *Blastomyces* cultures and primary specimens confirmed 43 cases of blastomycosis from NY from 2005 to 2019. These results further confirmed the endemicity of blastomycosis in New York (14). Of interest was the identification of five of 43 blastomycosis cases due to *B. gilchristii*. Interestingly, patients infected with *B. gilchristii* resided in different parts of NY, indicating broader geographic distribution of this species in NY. Additional studies are needed to determine if these cases are due to patients traveling between regions or if there is a focal niche of *B. gilchristii* in NY. Preponderance of blastomycosis cases due to *B. dermatitidis* with concentration in Mohawk Valley and Capital District followed by Finger Lakes, and sporadic cases from other regions of NY was noted. These results indicate that there is a broader distribution of *B. dermatitidis* compared to *B. gilchristii* in NY. These results are in agreement with earlier studies where *B. dermatitidis* has shown to have broader geographic range of blastomycosis cases from Canada to the southeastern United States while *B. gilchristii* appears limited to regions of Ontario, Saskatchewan, Alberta, Minnesota, and Wisconsin (25). The limited soil samples investigated in the present study from Mohawk Valley and Capital District failed to yield any positive DNA or culture of *B. dermatitidis* or *B. gilchristii*. These results are not surprising as the precise ecological niche(s) of *B. dermatitidis* or *B. gilchristii* is far from clear except in a few instance such as the Eagle River outbreak (26). An understanding of the ecological factors that favors growth, reproduction, and dispersal would potentially provide a means of predicting and controlling human acquisition of infection. This is particularly critical as in recent years, cases of blastomycosis in NY have increased and majority of patients reported no travel to regions of known endemicity (14). Given the seriousness of blastomycosis and consistently elevated incidence in NY, there is a proposal to make blastomycosis a reportable disease in NY.

Difference in virulence have long been observed among *Blastomyces* spp.(20, 27). It has been demonstrated previously that African strains of *B. dermatitidis* differ from North American strains in their growth and morphology and are thought to cause less severe disease (20). *BAD1* has been implicated as the major virulence factor of *B. dermatitidis* (28). The significant size difference in *BAD1* promoter identified in *B. gilchristii* and sequence analysis revealed that it was due to two large insertion (19). Whether these insertions have any influence on *BAD1* gene expression and virulence, need further investigation. Additionally, to the best of our knowledge, no studies are available to demonstrate the comparative virulence of *B. dermatitidis* and *B. gilchristii*. A number of cases of blastomycosis due to *B. gilchristii* including the fatal case of acute respiratory distress syndrome indicate that *B. gilchristii* pathogenic potential is comparable to that of *B. dermatitidis (29)*. A highly sensitive and specific assay would allow more comprehensive surveillance to monitor the disease incidence. Mandatory disease reporting and surveillance will also aid in the diagnosis of unknown cases, enable prompt initiation of treatment to decrease illness and death, and provide support for targeted public health interventions, such as public awareness campaigns (e.g., health advisories for blastomycosis in New York). In summary, the newly developed duplex real-time PCR assay would help understand the ecology and epidemiology of blastomycosis caused by *B. dermatitidis* and *B. gilchristii* in New York and North America.

## Data Availability

Yes

## ACKNOWLEDGMENTS

We thank Drs. Robert McDonald and Elizabeth Dufort, Division of Epidemiology, NYSDOH, and Elis Tobin, Albany Medical Center, Albany for providing soil samples and clinical specimens. We thank Wadsworth Center, Tissue Culture & Media, Histopathology, and Applied Genomic Technologies Cores for providing media, fixed tissue sectioning, and Sanger sequencing, respectively. This work was supported partly by the funds from the Wadsworth Center, New York State Department of Health (NYSDOH), and Centers for the Disease Control and Prevention (CDC) grant number NU50CK000516. In addition, Mitchell Kaplan was partially supported with National Science Foundation funding in the Wadsworth Center’s Research Experience for Undergraduates (REU) program (grant DBI1757170). Its contents are solely the responsibility of the authors and do not necessarily represent the official views of the Department of Health and Human Services or the CDC.

## AUTHOR CONTRIBUTIONS

SC conceived the study, supervised experiments and wrote the manuscript. MK designed primers and probes under YZ’s supervision and performed majority of the experiments, prepared graphs and tables. YZ supervised MK’s work, performed few key experiments, and prepared graphs and tables.VC edited the draft manuscript, JVK, and LM provided strains of *Blastomyces dermatitidis* and *B. gilchristii* and edited the draft manuscript.

---

**Supplementary Figure 1.**
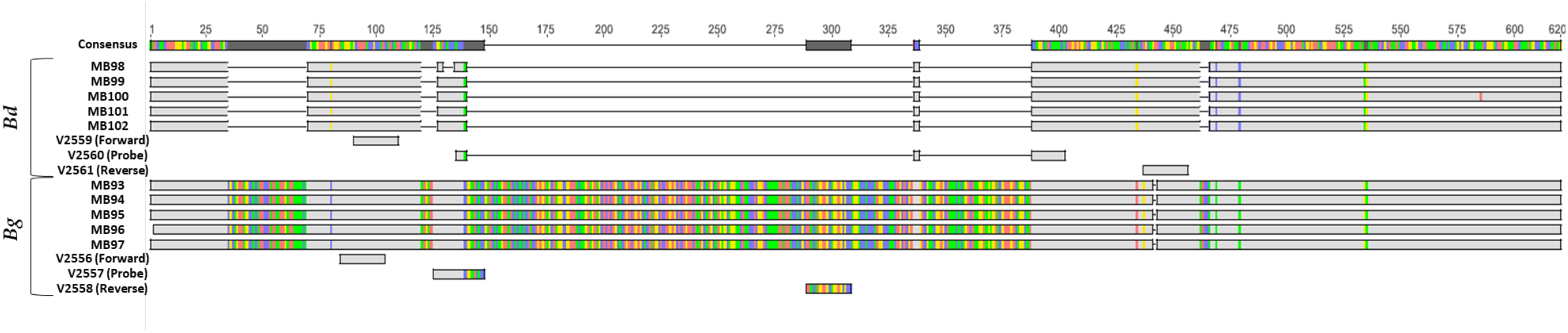
Multiple alignment of *BAD1* gene of *B. dermatitidis* and *B. gilchristii*. The sequences of *BAD1* gene from *B. dermatitidis* (Bd) and *B. gilchristii* (Bg) were aligned using MUSCLE alignment program in Geneious. The position of primers and probes sequences used in this study are shown.

**Supplementary Figure 2.**
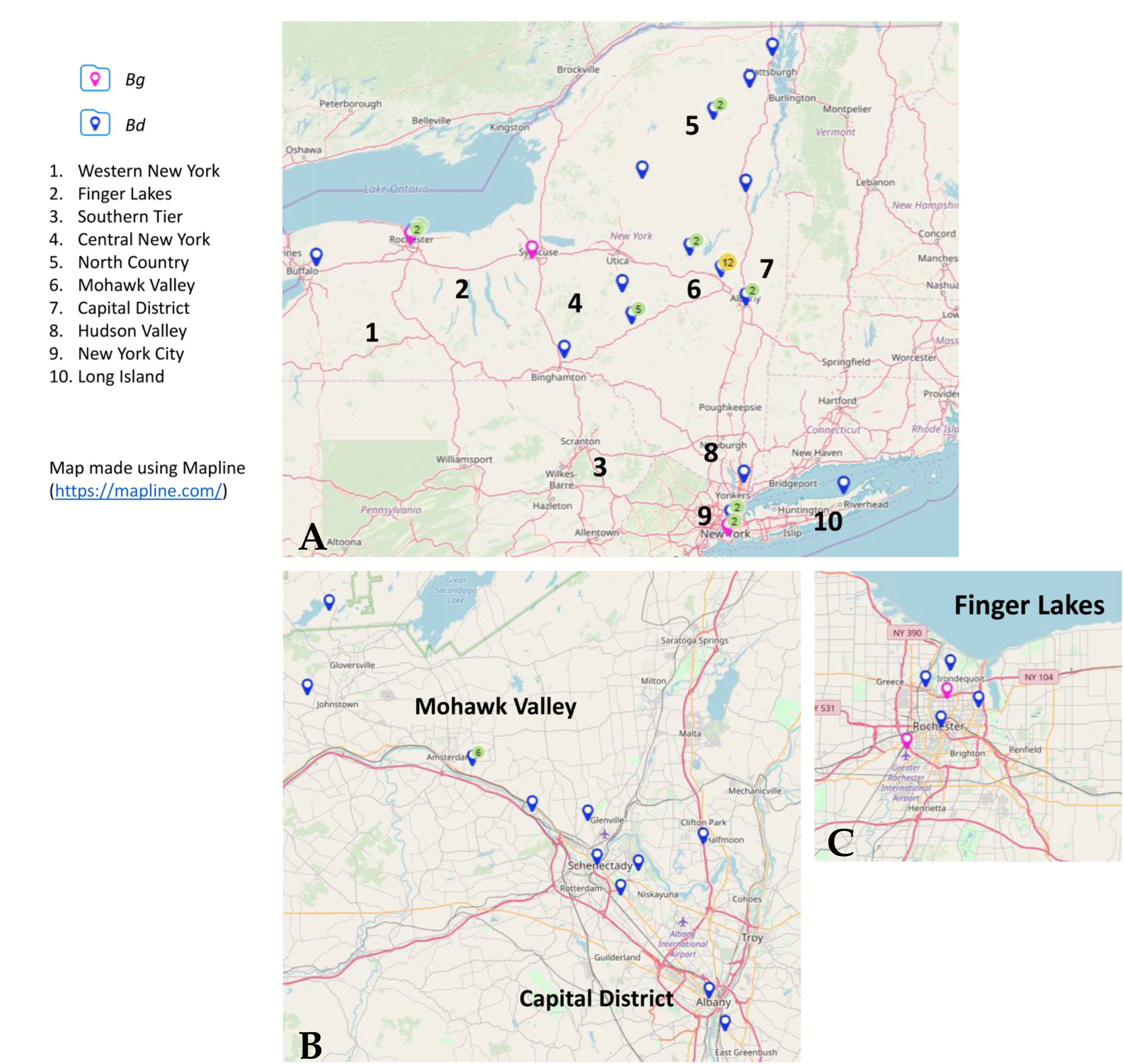
Geographic distribution of blastomycosis cases in New York from 2005 to 2019. (A) Patient’s residence (Zip Code) was utilized to map cases of blastomycosis due to *B. dermatitidis (Bd)* and *B. gilchristii (Bg)* in New York. (B) Clustering of cases in Mohawk Valley and Capital District, and (C) Finger Lakes, are shown.

**Supplementary Table 1.** Details of duplex real-time PCR assay of Blastomycosis cases from New York (2005-2019)

| Case No. | Primary Sample Collection Date | Source *Blastomyces* recovered in culture | Single plex real-time PCR (*Blastomyces* spp.) Ct (FAM) | Duplex real-time PCR |  | Final ID |
| --- | --- | --- | --- | --- | --- | --- |
|  |  |  |  | *Bd* Mean Ct (Cy5) | *Bg* Mean Ct (FAM) |  |
| Case 1 | 7/14/2005 | Bone | 22 | 22.03 | Undet | *B. dermatitidis* |
| Case 2 | 2/23/2006 | BAL | 20.7 | 20.71 | Undet | *B. dermatitidis* |
| Case 3 | 8/10/2006 | Tissue | 20.7 | 22.57 | Undet | *B. dermatitidis* |
| Case 4 | 1/17/2008 | Tissue | 22.5 | 22.57 | Undet | *B. dermatitidis* |
| Case 4 | 1/17/2008 | Tissue | 21.5 | 20.51 | Undet | *B. dermatitidis* |
| Case 4 | 1/17/2008 | Tissue | 21.2 | 20.46 | Undet | *B. dermatitidis* |
| Case 5 | 4/23/2008 | BAL | 20.19 | Undet | 20.67 | *B. gilchristii* |
| Case 6 | 8/17/2009 | Tissue | 22.01 | 21.31 | Undet | *B. dermatitidis* |
| Case 7 | 11/18/2009 | Tissue | Culture** | 24.17 | Undet | *B. dermatitidis* |
| Case 8 | 2/10/2010 | LUNG | 22.29 | Undet | 22.75 | *B. gilchristii* |
| Case 9 | 3/1/2012 | BAL | 23.33 | 22.43 | Undet | *B. dermatitidis* |
| Case 10 | 9/26/2012 | Lung | 20.97 | 19.47 | Undet | *B. dermatitidis* |
| Case 11 | 6/6/2013 | Aspirate | 20.94 | 18.57 | Undet | *B. dermatitidis* |
| Case 12 | 10/25/2013 | Bone | 18.8 | 23.58 | Undet | *B. dermatitidis* |
| Case 13 | 11/19/2013 | BAL | 20.54 | 22.12 | Undet | *B. dermatitidis* |
| Case 14 | 2/11/2014 | Skin lesion fluid* | 28.25 | 20.23 | Undet | *B. dermatitidis* |
| Case 15 | 2/17/2014 | Sputum | 20.52 | 22.47 | Undet | *B. dermatitidis* |
| Case 15 | 2/18/2014 | BAL | 24.34 | 21.46 | Undet | *B. dermatitidis* |
| Case 16 | 5/29/2014 | BAL | 27.63 | 20.73 | Undet | *B. dermatitidis* |
| Case 16 | 6/12/2014 | BAL | 21.73 | 18.99 | Undet | *B. dermatitidis* |
| Case 17 | 8/8/2014 | Brain | 28.99 | 27.95 | Undet | *B. dermatitidis* |
| Case 18 | 9/29/2015 | Cyst | 26.94 | 24.04 | Undet | *B. dermatitidis* |
| Case 19 | 10/20/2016 | Hand | 27.7 | 20.68 | Undet | *B. dermatitidis* |
| Case 20 | 1/13/2017 | Lung | 24.36 | 20.14 | Undet | *B. dermatitidis* |
| Case 21 | 1/16/2017 | BAL | 26.33 | Undet | 19.26 | *B. gilchristii* |
| Case 22 | 3/1/2017 | Lung | 26.79 | 23.74 | Undet | *B. dermatitidis* |
| Case 23 | 5/2/2017 | Sputum | 21.73 | 21.07 | Undet | *B. dermatitidis* |
| Case 24 | 5/17/2017 | BAL | 22.01 | 20.37 | Undet | *B. dermatitidis* |
| Case 25 | 6/28/2017 | Tissue | 23.22 | 23.06 | Undet | *B. dermatitidis* |
| Case 26 | 10/26/2017 | BAL | 21.19 | 23.49 | Undet | *B. dermatitidis* |
| Case 26 | 10/26/2017 | BAL | 21.34 | 24.37 | Undet | *B. dermatitidis* |
| Case 26 | 10/27/2017 | Sputum | 32.47 | 36.56 | Undet | *B. dermatitidis* |
| Case 26 | 10/27/2017 | Sputum | 29.71 | 32.02 | Undet | *B. dermatitidis* |
| Case 26 | 10/27/2017 | Sputum | 26.8 | 29.58 | Undet | *B. dermatitidis* |
| Case 27 | 10/3/2018 | Bone - Left Tibia | 19.11 | 21.54 | Undet | *B. dermatitidis* |
| Case 28 | 2/13/2018 | Bone (Tibia) | 20.05 | 22.71 | Undet | *B. dermatitidis* |
| Case 29 | 3/6/2018 | Tissue block - Left nasal Mass* | 37.4 | 39.63 | Undet | *B. dermatitidis* |
| Case 29 | 3/6/2018 | Tissue block - Left nasal Mass* | 33.29 | 37.24 | Undet | *B. dermatitidis* |
| Case 30 | 2/28/2018 | Tissue - Left Thigh | 20.47 | 23.58 | Undet | *B. dermatitidis* |
| Case 31 | 8/5/2018 | BAL | 22.63 | 27.74 | Undet | *B. dermatitidis* |
| Case 32 | 7/2/2018 | BAL | 22.89 | 27.36 | Undet | *B. dermatitidis* |
| Case 33 | 9/27/2018 | Tissue block* | 36.02 | 40.29 | Undet | *B. dermatitidis* |
| Case 34 | 10/2/2018 | Bone (Femur) | 19.09 | 22.77 | Undet | *B. dermatitidis* |
| Case 35 | 12/17/2018 | BAL | 19.72 | Undet | 22.49 | *B. gilchristii* |
| Case 36 | 1/20/2019 | Tissue | 20.87 | 24.81 | Undet | *B. dermatitidis* |
| Case 37 | 4/25/2019 | BAL | 21.01 | 24.54 | Undet | *B. dermatitidis* |
| Case 38 | 5/5/2019 | BAL* | 23.54 | 27.13 | Undet | *B. dermatitidis* |
| Case 39 | 5/9/2019 | BAL | 23.8 | 24.52 | Undet | *B. dermatitidis* |
| Case 40 | 5/23/2019 | Wound fluid | 20.16 | 23.26 | Undet | *B. dermatitidis* |
| Case 41 | 7/2/2019 | BAL | 18.74 | 23.06 | Undet | *B. dermatitidis* |
| Case 42 | 7/19/2019 | BAL | 19.45 | 24.28 | Undet | *B. dermatitidis* |
| Case 43 | 10/22/2019 | BAL | 18.88 | Undet | 17.73 | *B. gilchristii* |
| Case 43 | 10/22/2019 | BAL | 18.74 | Undet | 17.73 | *B. gilchristii* |

\*Primary samples; \*\* culture was confirmed by converting to the yeast form

**Supplementary Table 2.**
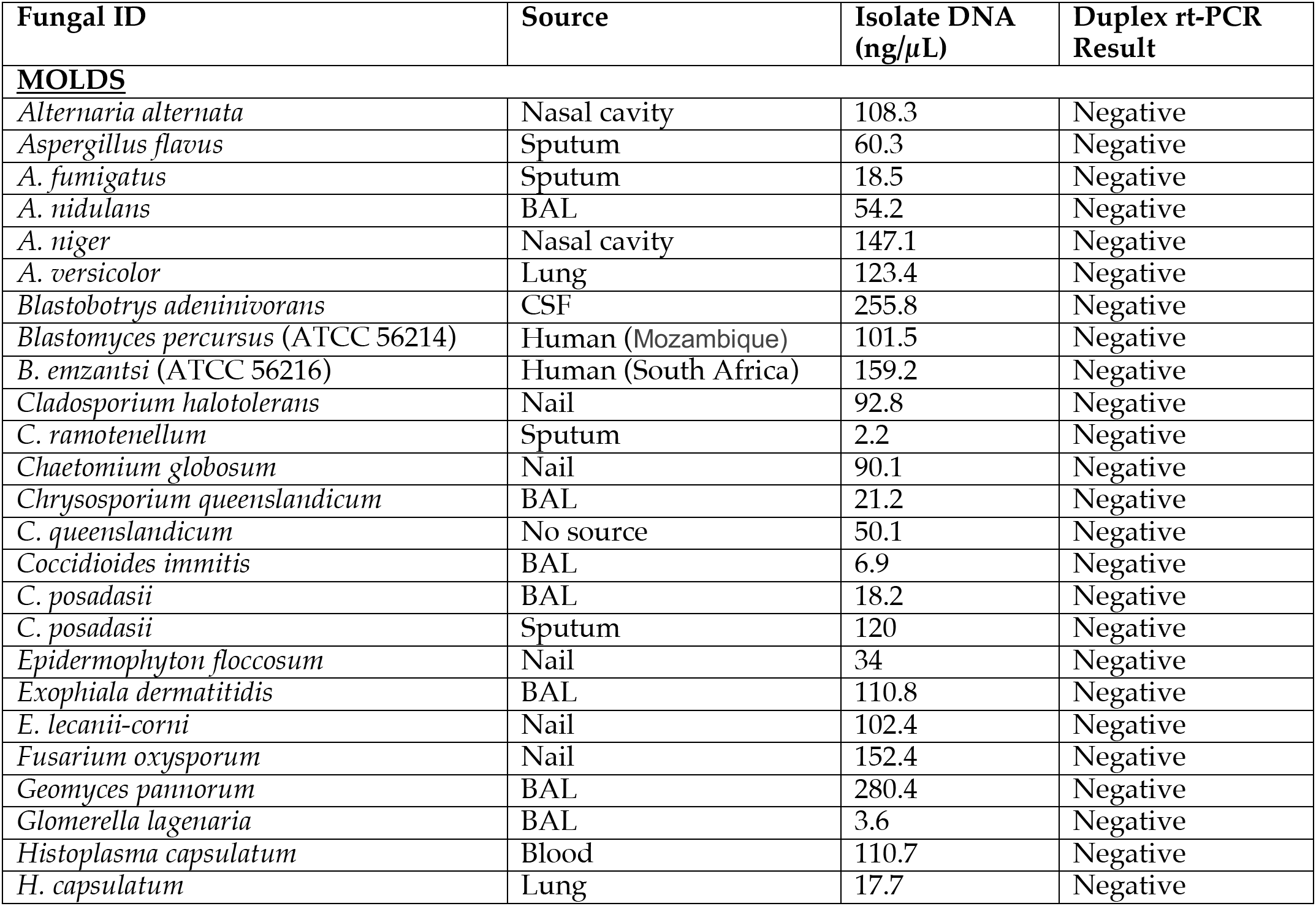
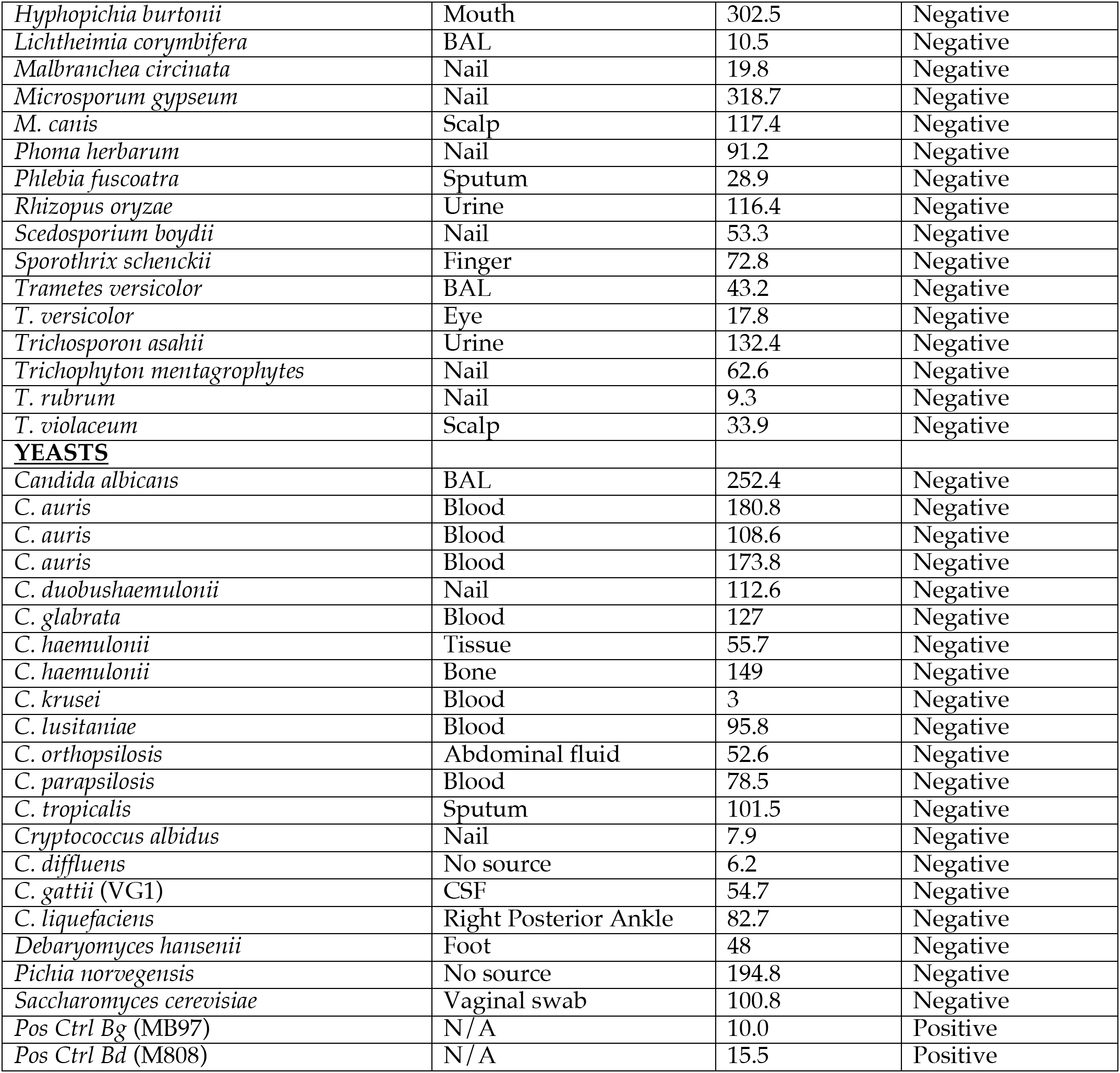
Specificity study of duplex real-time PCR assay

**Supplementary Table 3 A.**
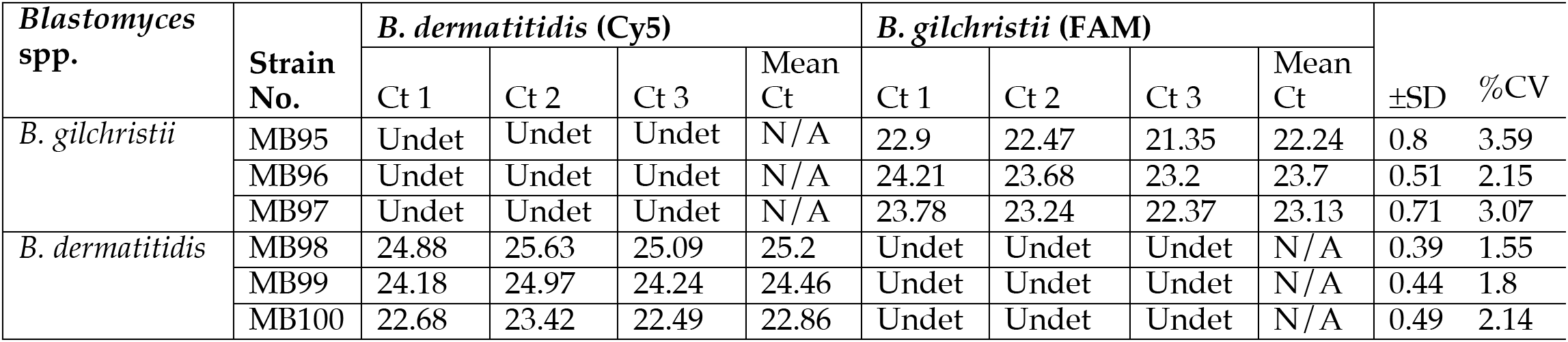
Inter-assay reproducibility of duplex real-time PCR assay

**Supplementary Table 3 B.**
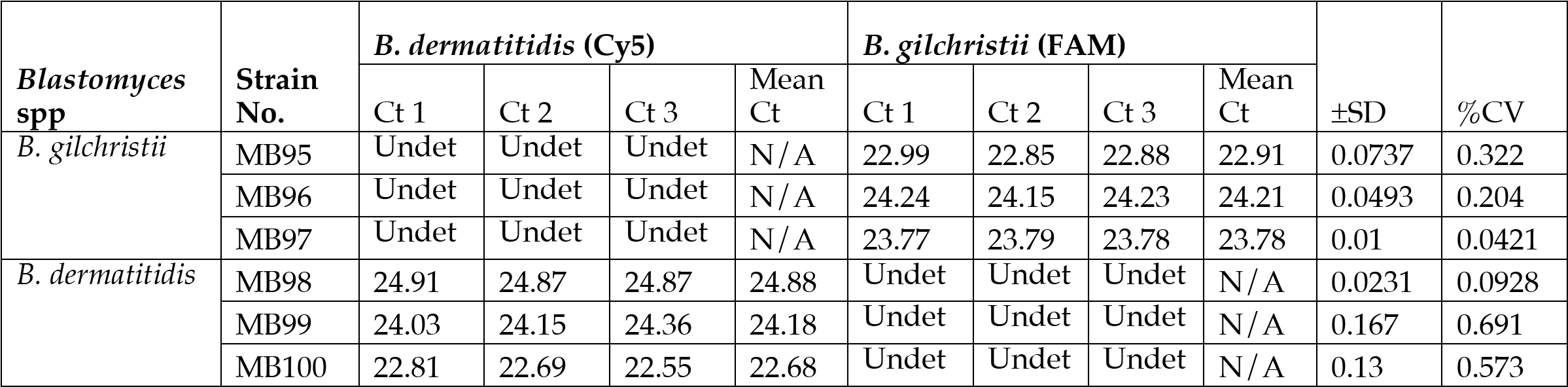
Intra-assay reproducibility of duplex real-time PCR assay

